# Drug exposure during oxycodone–S-ketamine patient-controlled analgesia reflects pain-related dosing behaviour after lumbar fusion surgery: a secondary analysis of a randomized clinical trial

**DOI:** 10.64898/2026.10.20.26365979

**Authors:** Taru Virtanen, Elina C.V. Brinck, Marica Engström, Mika J. Kurkela, Pekka Tarkkila, Janne Backman, Aleksi Tornio, Vesa Kontinen, Marko Peltoniemi, Teijo I. Saari

**Author notes:** **Correspondence**: Teijo Saari, University of Turku, Department of Anaesthesiology and Intensive Care, P.O. Box 52 (Kiinamyllynkatu 4-8, 11A5), FI-20521 Turku, Finland. **Clinical Trial Number:** NCT02994173, https://clinicaltrials.gov/study/NCT02994173.

## Abstract

**Background:** We examined the relationships between acute pain intensity, patient-controlled dosing behaviour, and drug exposure during postoperative oxycodone patient-controlled analgesia (PCA) with or without adjunctive S-ketamine.

**Methods:** This observational secondary analysis included 100 patients undergoing lumbar fusion surgery in a randomized, double-blind trial comparing oxycodone PCA alone with three concentrations of adjunctive S-ketamine.Plasma concentration data were available for 99 patients. Generalized estimating equations were used to examine dosing behaviour, drug exposure, and adequate analgesia (pain intensity at rest ≤4/10) during the first 24 h.

**Results:** Plasma concentrations of oxycodone and S-ketamine increased rapidly after PCA initiation, with substantial between-patient variability. Metabolite concentrations accumulated over time. PCA activity decreased over time, and older age was associated with fewer delivered doses (rate ratio [RR] 0.905 per 10 years, 95% CI 0.829–0.987). Each 1-point higher current pain score was associated with more subsequent PCA attempts (RR 1.09, 95% CI 1.04–1.14) and delivered doses (RR 1.07, 95% CI 1.03–1.11). The association between oxycodone concentration and adequate analgesia weakened over time (interaction P < 0.001), with odds ratios per 10 ng/mL of 1.16 (95% CI 1.07–1.25), 1.13 (1.06–1.21), and 1.00 (0.979–1.03) at 1, 4, and 24 h, respectively. In S-ketamine-treated patients, these associations persisted after adjustment for measured S-ketamine and metabolite concentrations.

**Conclusions:** During PCA, drug exposure develops from patient-generated dosing histories while pain and recovery change. Pain intensity, patient-controlled dosing behaviour, and drug exposure are interconnected but non-equivalent features of postoperative analgesia.

**What is already known on this topic:** Opioid consumption during patient-controlled analgesia is commonly used as an outcome in postoperative analgesia trials, but its relationship with pain intensity and systemic drug exposure over time is incompletely understood.

**What this study adds:** Higher current pain was associated with greater subsequent patient-controlled dosing activity. The association between plasma oxycodone concentration and adequate analgesia weakened during the first 24 postoperative hours. Achieved S-ketamine exposure reflected both randomized treatment allocation and patient-controlled dosing behaviour.

**How this study might affect research, practice or policy:** Pain intensity, opioid consumption, and systemic drug exposure should be considered distinct but interrelated outcomes when interpreting postoperative patient-controlled analgesia studies.

## INTRODUCTION

Opioid patient-controlled analgesia (PCA) allows patients to self-administer intravenous opioids within predefined safety limits^1^. Compared with conventional systemic opioid administration, PCA provides slightly better pain control and greater patient satisfaction, although opioid consumption may be higher^2^. Intravenous oxycodone PCA may provide effective analgesia at lower doses than morphine in some surgical settings^3,4^.

PCA differs fundamentally from a predetermined dosing regimen because the patient participates in creating the dose history^1,2^. PCA requirements vary with pain state and patient-related factors^5,6^, while successful requests generate delivered opioid doses from which cumulative administration and systemic exposure develop over time^3,4,7^. We therefore conceptualized PCA as a patient-responsive titration process in which pain state, dosing behaviour, administered drug, and exposure evolve together over time. This framework specifies temporal relationships rather than an automated or causal feedback system.

The pharmacokinetics of oxycodone and its relationship with analgesic effects have been characterized in several clinical settings^7,8^. Plasma oxycodone concentration is a pharmacologically relevant measure of exposure, but its interpretation during PCA is not straightforward. A concentration measured at a given postoperative time reflects the preceding dosing history, while the clinical state in which that concentration is observed is also evolving. Consequently, the association between plasma oxycodone concentration and pain during PCA needs to be considered in the temporal context in which both exposure and postoperative pain are evolving. How this relationship changes during the early course of postoperative PCA has not been well characterized.

Systematic reviews have shown opioid-sparing effects of adjunctive ketamine, including in patients undergoing spinal surgery^9,10,11^. In our randomized trial after lumbar fusion surgery, adjunctive S-ketamine reduced oxycodone consumption dose-dependently, with modest improvement in pain at rest at the highest dose^12^. During PCA, however, achieved S-ketamine exposure depends on both randomized treatment allocation and patient-controlled dosing.

Age has been associated with postoperative PCA requirements^5,6^ as well as oxycodone clearance and systemic exposure^13,14,15^. In our lumbar fusion cohort, preoperative emotional stress was associated with severe postoperative pain but not with oxycodone consumption^16^. Together, these observations suggest that pain intensity, opioid requirements, and systemic drug exposure need not vary in parallel.

The primary objectives of this analysis were to examine the association between current pain and subsequent PCA activity and whether the association between plasma oxycodone concentration and adequate analgesia varied over time. We also characterized drug exposure and age-related PCA behaviour during the first 24 h and assessed the robustness of the primary associations to adjustment for S-ketamine and metabolite concentrations.

## METHODS

### Study design

We conducted an observational secondary analysis of the DoseRespKeta study, a randomized, double-blind trial comparing postoperative PCA with oxycodone alone or combined with S-ketamine at three dose ratios after lumbar fusion surgery. The trial protocol, eligibility criteria, and primary results have been published previously.¹² Blood sampling and plasma drug concentration measurements were included in the approved study protocol, but the present analyses were not prespecified. The study was approved by the institutional review board of the Hospital District of Southwest Finland (103/1800/2016) and the Finnish Medicines Agency (FIMEA, KL 135/2016). The study was registered before patient enrolment at ClinicalTrials.gov (NCT02994173; https://clinicaltrials.gov/study/NCT02994173; principal investigator: Teijo I. Saari; registration date: November 11, 2016) and in the EudraCT database (2016-002887-14; principal investigator: Teijo I. Saari; registration date: October 12, 2016). The trial was conducted between February 2017 and May 2021 at Turku University Hospital and Helsinki University Hospital. First patient was recruited on 6 February 2017. This observational secondary analysis is reported in accordance with the Strengthening the Reporting of Observational Studies in Epidemiology (STROBE) guidelines^17^.

### Randomization and postoperative analgesia and PCA protocol

Participants were randomized 1:1:1:1 to PCA with oxycodone 1 mg/mL alone (G1) or combined with S-ketamine 0.25 mg/mL (G2), 0.5 mg/mL (G3), or 0.75 mg/mL (G4). Randomization, masking, anaesthesia, and perioperative care have been described in detail previously^12^. Treatment group was retained as an adjustment variable in the present analyses; parent oxycodone was the principal exposure analyte.

The PCA pump was activated at the end of surgery. In the post-anaesthesia care unit, the study solution was titrated in 2-mL doses at 5-min intervals until pain at rest was ≤4 on the numerical rating scale (NRSR). Thereafter, each successful PCA request delivered 1 mL with a 5-min lockout. The randomized study solution was continued for 24 h^12^.

### Pain assessment

Pain intensity at rest was assessed using an 11-point numerical rating scale (NRSR; 0 = no pain and 10 = worst pain imaginable). Pain assessments were performed according to a predefined schedule as part of postoperative care, with additional assessments obtained at the time of blood sampling. Adequate analgesia was defined as NRSR ≤4.

### Blood sampling and bioanalysis

Arterial blood (4 mL) was collected through the intraoperative arterial cannula at baseline and at 5, 15, 30–60 min, 4 h, and 18–24 h after PCA initiation into EDTA tubes. Plasma was separated within 30 min, aliquoted, and stored at −70 °C until analysis.

Plasma concentrations of oxycodone, noroxycodone, oxymorphone, and noroxymorphone and of ketamine, norketamine, and 6-hydroxynorketamine were determined using validated HPLC– MS/MS methods. The ketamine assay was not enantioselective^18,19^. We administered only S-ketamine, and ketamine enantiomers do not undergo interconversion^19^. Thus, the measured ketamine represented S-ketamine. The lower limits of quantification were 0.05 ng/mL for oxycodone and its metabolites, 0.025 ng/mL for ketamine, 0.25 ng/mL for norketamine, and 0.5 ng/mL for 6-hydroxynorketamine. Full analytical methods are provided in the Supplementary Methods.

### Statistical analysis

The sample size was determined by the parent trial^12^ without additional sample size calculation. Analyses used available observations without imputation of missing data. Participants with missing plasma samples contributed to analyses not requiring concentration data.

Continuous variables are presented as mean (standard deviation) or median (interquartile range), and categorical variables as counts and percentages. Analyses were restricted to the first 24 h after PCA initiation. Drug concentrations were summarized descriptively at protocol sampling times.

Longitudinal associations of time and age with PCA attempts, delivered doses, cumulative oxycodone dose, and plasma oxycodone concentration were analysed using generalized estimating equations (GEEs), adjusting for randomized treatment group. Models included time, age, and their interaction. Count outcomes used Poisson GEEs with log interval duration as an offset and continuous outcomes identity-link GEEs. Age was centred at the sample median; tertiles were used only for visualization. Exploratory models replaced age with body weight.

The relationship between current pain and subsequent PCA activity was examined over intervals between consecutive observed NRSR assessments. PCA attempts and delivered doses during each subsequent interval were analysed using Poisson GEEs including current NRSR, time, age, and treatment group, with log interval duration as an offset.

The association between plasma oxycodone concentration and adequate analgesia (NRSR ≤4) was analysed using logistic GEE including concentration, NRSR assessment time, their interaction, age, and treatment group. Concentrations were paired with simultaneous NRSR assessments or, if unavailable, the nearest assessment within ±15 min; each concentration was used once. Time-specific odds ratios were estimated per 10 ng/mL higher oxycodone concentration.

Sensitivity analyses in the active S-ketamine groups (G2–G4) examined whether these associations were robust to measured S-ketamine and metabolite exposure. Analyses used the 15-, 60-, and 240-min epochs for pain–PCA models and additionally 1440 min for oxycodone– analgesia models. Log2-transformed S-ketamine, norketamine, and 6-hydroxynorketamine concentrations were centred within treatment group and sampling epoch. The original model was refitted on the common subset (M0), followed by addition of S-ketamine (M1) and all three analytes (M2). Additional sensitivity analyses excluded the 15-min sampling epoch or observations with any S-ketamine or metabolite concentration below the limit of quantification. As measured concentrations are post-randomization exposures influenced by preceding PCA use, their coefficients were interpreted as associations rather than causal effects.

All GEEs accounted for within-patient clustering using robust sandwich standard errors. Independence working correlation structures were used for longitudinal age and weight analyses, and exchangeable structures for pain–PCA, oxycodone–analgesia, and S-ketamine-adjusted analyses.

Associations are reported as effect estimates with 95% confidence intervals. Statistical tests were two-sided, with a significance level of 0.05 and no adjustment for multiple comparisons. Analyses used R version 4.5.2 (R Foundation for Statistical Computing, Vienna, Austria), with *tidyverse* and *ggplot2* for data management and visualization and *geepack* for GEE modelling.

## RESULTS

### Study population

Oxycodone and metabolite concentration data were available for 99 participants (586 observations per analyte; 92 complete profiles). Other analyses used available data from the original 100 participants. Participant flow and analysis populations are shown in Supplementary Figure S1. Analysis-specific sample sizes are reported in the corresponding tables. Baseline characteristics are shown in Table 1.

**Table 1.** Baseline characteristics of the study population according to randomized treatment group.

| **Characteristic** | **All** | **G1:**<br>Oxycodone 1 mg/mL | **G2:**<br>Oxycodone 1 mg/mL +<br>S-ketamine 0.25 mg/mL | **G3:**<br>Oxycodone 1 mg/mL +<br>S-ketamine 0.5 mg/mL | **G4:**<br>Oxycodone 1 mg/mL +<br>S-ketamine 0.75 mg/mL |
|---|---|---|---|---|---|
| Sample size, n | 99 | 26 | 25 | 24 | 24 |
| Age, yr, mean (SD) | 60.8 (11.5) | 59.2 (13.0) | 62.2 (8.3) | 58.9 (12.4) | 62.9 (11.8) |
| Weight, kg, mean (SD) | 77.7 (15.4) | 76.3 (14.8) | 80.1 (15.4) | 77.8 (13.4) | 76.4 (18.3) |
| Sex, n (%) |  |  |  |  |  |
| Female | 65 (66) | 18 (69) | 16 (64) | 15 (63) | 16 (67) |
| Male | 34 (34) | 8 (31) | 9 (36) | 9 (37) | 8 (33) |
| Previous chronic pain, n (%) |  |  |  |  |  |
| Yes | 73 (74) | 18 (69) | 16 (64) | 21 (88) | 18 (75) |
| No | 26 (26) | 8 (31) | 9 (36) | 3 (13) | 6 (25) |
| Preoperative opioid use, n (%) |  |  |  |  |  |
| Yes | 10 (10) | 3 (12) | 3 (12) | 2 (8.3) | 2 (8.3) |
| No | 89 (90) | 23 (88) | 22 (88) | 22 (92) | 22 (92) |
| Preoperative pain on movement, (median, IQR) | 4 (2-7) | 4 (3-7) | 4 (2-7) | 4.5 (3-5) | 4 (0-7) |

Baseline characteristics are presented for the 99 participants with available plasma oxycodone concentration data. IQR, interquartile range; SD, standard deviation.

### Oxycodone exposure during PCA

Plasma oxycodone concentrations increased rapidly after PCA initiation, with substantial between-patient variability. Noroxycodone, oxymorphone, and noroxymorphone concentrations were initially low and increased over subsequent hours (Figure 1A). Profiles stratified by randomized treatment group are shown in Supplementary Figure S2.

**Figure 1.**
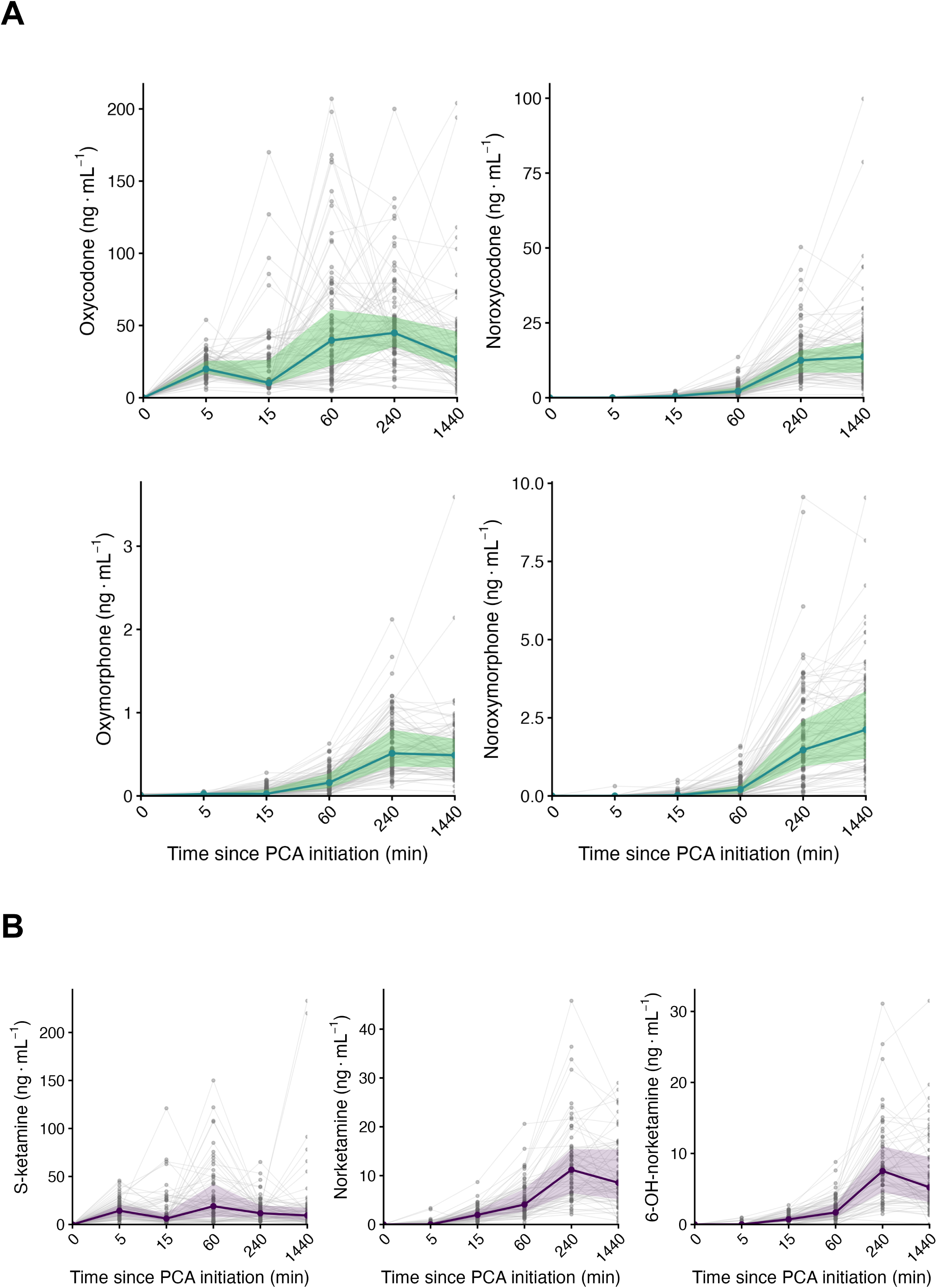
Drug exposure during the first 24 h of oxycodone–S-ketamine patient-controlled analgesia (PCA). (A) Plasma concentrations of oxycodone, noroxycodone, oxymorphone, and noroxymorphone at protocol sampling epochs after PCA initiation. (B) Plasma concentrations of S-ketamine, norketamine, and 6-hydroxynorketamine in participants randomized to S-ketamine-containing PCA. Grey lines and points show individual concentration trajectories and observations; coloured lines and ribbons show the median and interquartile range. Protocol sampling epochs are displayed at equal spacing. The three highest oxycodone concentrations and the highest S-ketamine concentration at 24 h were omitted from the display of individual observations to improve visualization but were retained in all summary calculations and analyses.

### S-ketamine and metabolite exposure during PCA

S-ketamine was measurable early after PCA initiation, whereas norketamine and 6-hydroxynorketamine concentrations increased more gradually (Figure 1B). Concentration profiles stratified by randomized treatment group are shown in Supplementary Figure S3. At 5 min, 6-hydroxynorketamine was quantifiable in only 6 participants. Cumulative S-ketamine dose varied both between and within randomized treatment groups and at 24 h, median (IQR) cumulative doses were 17.2 (12.8–23.2), 25.0 (20.8–44.3), and 54.0 (35.8–69.9) mg in G2, G3, and G4, respectively.

Realized cumulative S-ketamine dose accounted for more of the variability in log2-transformed plasma S-ketamine concentration than randomized treatment group at 15, 60, and 240 min (R² 0.490 vs 0.346, 0.357 vs 0.172, and 0.623 vs 0.301, respectively), whereas the difference was small at 24 h (0.186 vs 0.169).

### Patient-controlled dosing behaviour and exposure

The longitudinal PCA activity analyses included 4529 intervals from 96 participants. PCA attempt and delivered-dose rates decreased with postoperative time (RR 0.965 and 0.966 per hour, respectively; both P<0.001; Supplementary Table S2 and Figure 2). Older age was associated with a lower delivered-dose rate (RR 0.905 per 10 years, 95% CI 0.829–0.987; P=0.02), whereas the association with attempt rate was less precise (RR 0.904, 95% CI 0.805– 1.02; P=0.09). There was no evidence of age-by-time interactions for either outcome.

**Figure 2.**
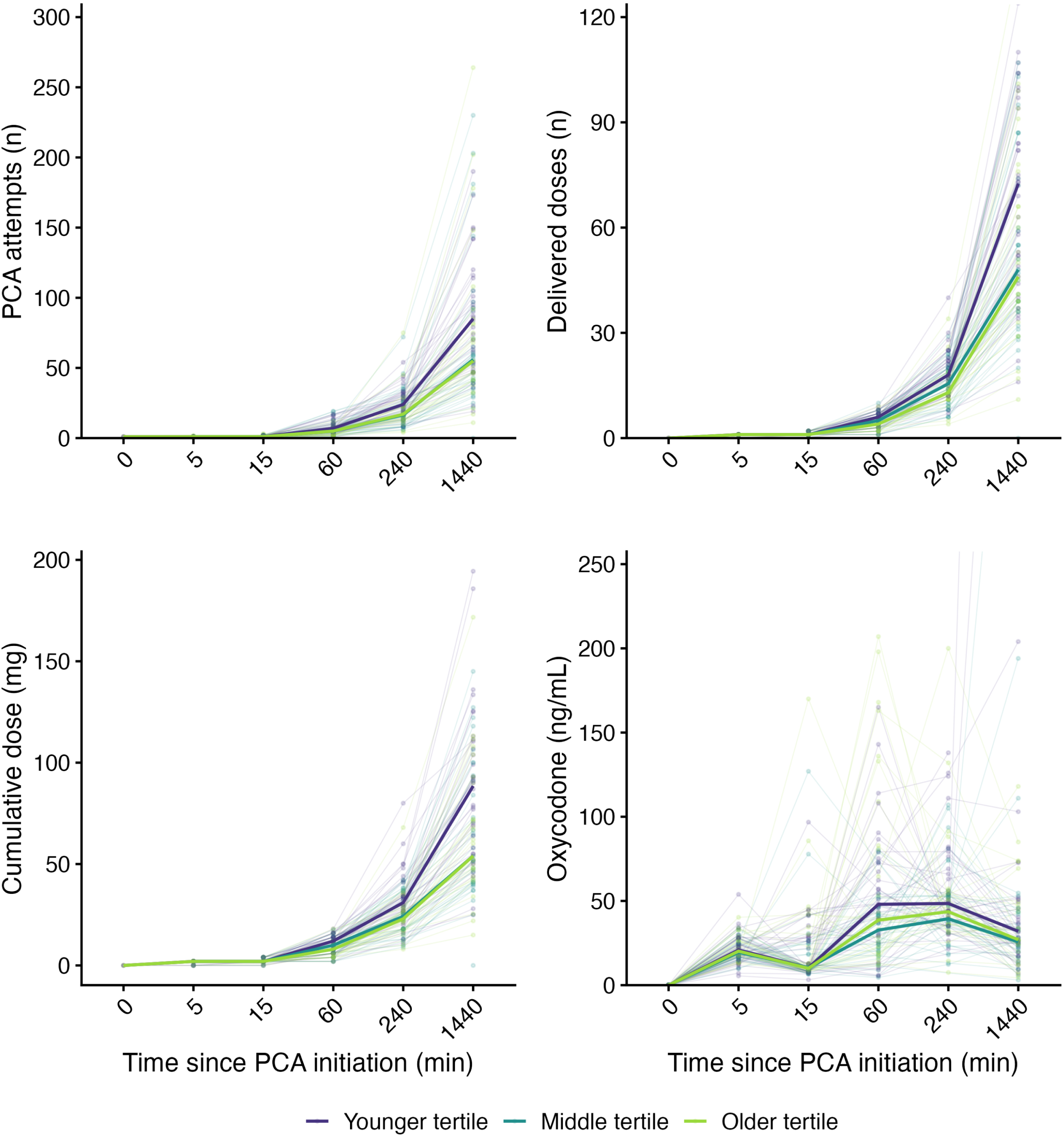
Patient-controlled dosing behaviour and exposure over time according to age. Individual trajectories and age-stratified summaries are shown for (A) cumulative PCA attempts, (B) cumulative delivered PCA doses, (C) cumulative oxycodone dose, and (D) plasma oxycodone concentration during the first 24 h after PCA initiation. Thin lines and points represent individual participants; thick coloured lines indicate medians. Protocol sampling epochs are displayed at equal spacing. Colour indicates age tertile: 30–58 years (n=35), 59–68 years (n=34), and 70–78 years (n=31). Age tertiles were used for visualization only; age was analysed continuously in the statistical models. PCA, patient-controlled analgesia.

Cumulative oxycodone dose increased by an estimated 3.03 mg/h (95% CI 2.69–3.37; P<0.001) at median age. The age-by-time interaction was −0.391 mg/h per 10-year increase in age (95% CI −0.818 to 0.0363; P=0.07). Plasma oxycodone concentration increased by an estimated 1.57 ng/mL/h (95% CI 0.346–2.79; P=0.01) at median age, with no evidence of an age-by-time interaction (Supplementary Table S2).

### Current pain and subsequent PCA activity

The analysis included 633 intervals between consecutive pain assessments in 100 participants. Higher current pain intensity was associated with greater PCA activity during the subsequent interval (Table 2 and Figure 3A–B). Each 1-point higher NRSR was associated with 9% more subsequent attempts (RR 1.09, 95% CI 1.04–1.14; *P*<0.001) and 7% more subsequent delivered doses (RR 1.07, 95% CI 1.03–1.11; *P*<0.001). Both rates decreased with postoperative time.

**Figure 3.**
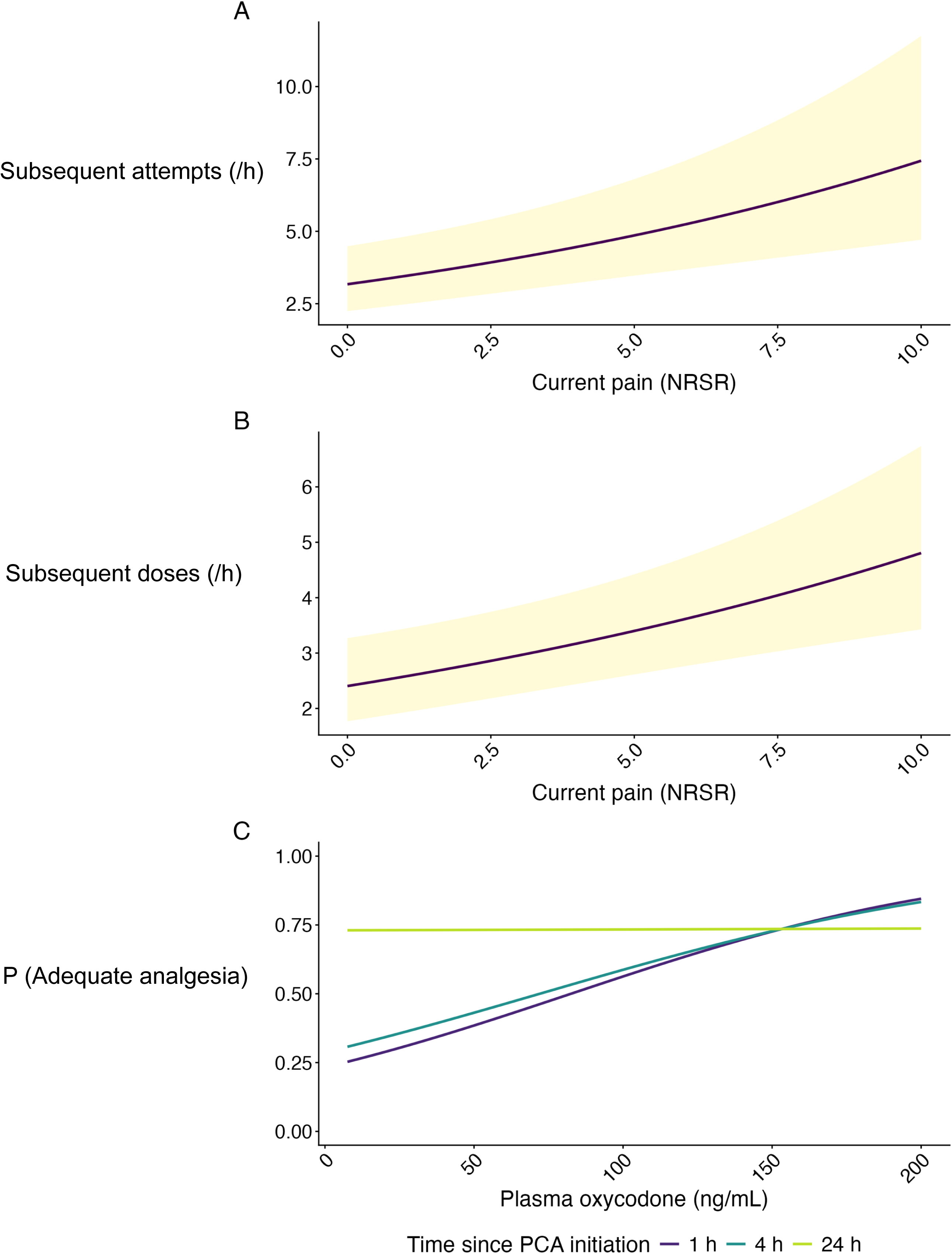
Model-based associations between current pain, subsequent PCA behaviour, and plasma oxycodone concentration during the first 24 h of patient-controlled analgesia. (A) Estimated hourly rate of subsequent PCA attempts and (B) estimated hourly rate of subsequent delivered PCA doses according to the pain score at rest (NRSR) at the beginning of each interval. Lines show model estimates and ribbons indicate 95% confidence intervals. Predictions are shown at median postoperative time and age for the oxycodone-only treatment group (G1). (C) Model-based probability of adequate analgesia (NRSR ≤4) according to plasma oxycodone concentration at 1, 4, and 24 h after PCA initiation. Predictions are shown at median age for G1. NRSR, numerical rating scale pain score at rest; PCA, patient-controlled analgesia.

**Table 2.** Associations of current pain intensity with subsequent PCA activity and of plasma oxycodone concentration with adequate analgesia during the first 24 h after PCA initiation.

| **Outcome** | **Model term or postoperative time** | **Estimate (95% CI)** | **P value** |
|---|---|---|---|
| Subsequent PCA attempts<sup>a</sup> | Current NRSR, per 1 point | 1.09 (1.04–1.14) | <0.001 |
|  | Postoperative time, per 1 h | 0.931 (0.905–0.958) | <0.001 |
|  | Age, per 10 years | 0.911 (0.796–1.04) | 0.18 |
| Subsequent delivered PCA doses<sup>a</sup> | Current NRSR, per 1 point | 1.07 (1.03–1.11) | <0.001 |
|  | Postoperative time, per 1 h | 0.933 (0.913–0.955) | <0.001 |
|  | Age, per 10 years | 0.891 (0.813–0.977) | 0.01 |
| Adequate analgesia<sup>b,c</sup><br>OR per 10 ng/mL increase in plasma oxycodone | 1 h | 1.16 (1.07–1.25) | — |
|  | 4 h | 1.13 (1.06–1.21) | — |
|  | 24 h | 1.00 (0.979–1.03) | — |

<sup>a</sup> PCA attempt and delivered-dose models used Poisson GEE with a log link and log interval duration as an offset. Estimates are reported as rate ratios (RRs). Current NRSR refers to pain intensity at the beginning of each interval, and PCA activity to events occurring during the subsequent interval. Models included current NRSR, postoperative time at interval start, age, and randomized treatment group. Both models included 633 intervals from 100 participants.
<sup>b</sup> Adequate analgesia was defined as NRSR ≤4. Plasma oxycodone concentrations were matched to simultaneous NRSR assessments or, when unavailable, to the nearest NRSR assessment within ±15 min. The logistic GEE model included plasma oxycodone concentration, postoperative time, their interaction, age, and randomized treatment group. Postoperative time was defined by the matched NRSR assessment. The model included 548 matched concentration–NRSR observations from 99 participants.
<sup>c</sup> The association between plasma oxycodone concentration and adequate analgesia varied with postoperative time (concentration-by-time interaction, P<0.001). Time-specific odds ratios (ORs) and 95% CIs were derived from linear contrasts of the fitted interaction model and represent the association with adequate analgesia per 10 ng/mL increase in plasma oxycodone concentration at 1 h , 4 h, and 24 h after PCA initiation. Individual P values are not reported for these contrasts.
All models used an exchangeable working correlation structure, participant-level clustering, and robust sandwich standard errors. P values are from two-sided Wald tests.
CI, confidence interval; GEE, generalized estimating equations; NRSR, numerical rating scale pain score at rest; OR, odds ratio; PCA, patient-controlled analgesia; RR, rate ratio.

Older age was associated with fewer subsequent delivered doses (RR 0.891 per 10 years, 95% CI 0.813–0.977; P=0.01), whereas the association with subsequent attempts was less precise (RR 0.911, 95% CI 0.796–1.04; P=0.18). In the ketamine-treated groups, the associations between current pain and subsequent PCA activity were essentially unchanged after adjustment for measured S-ketamine and metabolite concentrations; across M0–M2, the rate ratio per one-point higher NRSR was 1.08–1.09 for subsequent PCA attempts and 1.06–1.07 for delivered doses (Supplementary Table S3).

### Plasma oxycodone and adequate analgesia

Adequate analgesia was observed in 37/96 (38.5%), 77/97 (79.4%), and 66/95 (69.5%) patients at the 1-, 4-, and 24-h sampling epochs, respectively. The association between plasma oxycodone concentration and adequate analgesia varied with postoperative time (concentration-by-time interaction, P<0.001; Table 2 and Figure 3C). Per 10 ng/mL higher plasma oxycodone concentration, the odds ratio for adequate analgesia was 1.16 (95% CI 1.07–1.25) at 1 h and 1.13 (95% CI 1.06–1.21) at 4 h, compared with 1.00 (95% CI 0.979–1.03) at 24 h.

Among the ketamine-treated groups, this temporal pattern persisted after inclusion of measured S-ketamine, norketamine, and 6-hydroxynorketamine concentrations (Supplementary Table S3); in the fully adjusted model, the corresponding odds ratios were 1.30 (95% CI 1.10–1.53), 1.26 (1.09–1.45), and 1.02 (0.971–1.07) at 1, 4, and 24 h, respectively. Within this post-randomization exposure analysis, higher measured S-ketamine concentration was associated with lower odds of adequate analgesia, whereas no clear associations were observed for norketamine or 6-hydroxynorketamine (Supplementary Table S4). This association was not reproduced in the analysis restricted to the 60-, 240-, and 1440-min epochs (Supplementary Table S4).

### Exploratory analysis of body weight

Body weight showed no clear associations with PCA attempts, delivered doses, cumulative oxycodone dose, or plasma oxycodone concentration, and no weight-by-time interactions were observed (Supplementary Table S5).

## DISCUSSION

During postoperative patient-controlled analgesia, opioid exposure reflects the patient’s dosing history while pain and recovery change over time. Our results show that higher current pain was associated with greater subsequent PCA activity, whereas the association between plasma oxycodone concentration and adequate analgesia weakened with postoperative time. Achieved S-ketamine exposure reflected both randomized treatment allocation and patient-controlled dosing.

PCA allows patients to request doses in response to perceived need. Previous studies have described substantial variability in PCA requirements and identified patient-related factors associated with opioid use^1,2,5,6^. Our findings extend previous observations by showing a temporal association between pain and subsequent PCA use. Higher pain was associated with more subsequent PCA attempts and delivered doses. These associations remained essentially unchanged after adjustment for measured S-ketamine and metabolite concentrations. The temporal relationship is consistent with a patient-responsive titration process but does not establish a causal feedback relationship.

Age may influence postoperative oxycodone use through several mechanisms^5,6^. Previous studies of morphine PCA have reported lower opioid requirements with increasing age^20–22^. Oxycodone clearance decreases with advancing age, and age-related differences in systemic exposure have been reported in postoperative patients^13,14,15^. In the present longitudinal analyses, the association between age and PCA attempts was less certain. Furthermore, there was no clear evidence of an association between age and plasma oxycodone concentration over time. The clearest age association was observed for delivered PCA dosing, although our data do not distinguish between behavioural, pharmacological, and other possible explanations.

Pain intensity, patient-controlled opioid use, and plasma exposure should not be regarded as interchangeable measures of the postoperative state. Previous studies of morphine PCA have demonstrated that postoperative pain and opioid use are related, but their associations with patient characteristics do not necessarily coincide^22,23^. In our previous analysis of this cohort, preoperative emotional stress was associated with severe postoperative pain but not with oxycodone consumption^16^. In the present study, current pain was associated with subsequent PCA activity, showing that pain and patient-controlled dosing behaviour are related but distinct observations. The age analyses further indicated that associations with PCA attempts, delivered doses, and plasma oxycodone concentration were not uniform. These findings indicate that pain, patient-controlled dosing behaviour, and opioid exposure are related, but not interchangeable and need not vary in parallel.

S-ketamine concentrations increased early after PCA initiation, whereas norketamine and 6-hydroxynorketamine exposure built up more gradually. Given the pharmacological activity of these metabolites, their slower formation may have clinical relevance during the early postoperative period, when analgesic requirements are high^24–26^. Randomization determined the S-ketamine concentration in the PCA solution, but achieved exposure also depended on patient-controlled dosing. Indeed, cumulative S-ketamine dose explained more variability in plasma S-ketamine concentration than randomized treatment group during the early and intermediate sampling periods. Higher S-ketamine concentration was associated with lower odds of adequate analgesia, but this post-randomization association should not be interpreted as evidence of impaired analgesia. Greater analgesic need may lead to more PCA use, a higher S-ketamine dose, and consequently higher exposure. This interpretation is consistent with the parent trial, in which adjunctive S-ketamine reduced oxycodone consumption dose-dependently and the highest dose also modestly improved pain at rest^12^.

Previous clinical PK/PD studies have described a relationship between oxycodone exposure and analgesic effect^7,8,27^. Consistent with these findings, higher plasma oxycodone concentrations were associated with a greater probability of adequate analgesia early after PCA initiation. However, this association weakened with postoperative time and was no longer apparent at 24 h. The same temporal pattern persisted after inclusion of measured S-ketamine, norketamine, and 6-hydroxynorketamine concentrations. This attenuation does not imply that concentration is unimportant. Rather, our findings suggest that the relationship between plasma oxycodone concentration and analgesia changes over the course of postoperative PCA.

During PCA, postoperative time is closely linked to oxycodone exposure because the observed concentration reflects the dosing history accumulated up to that point. At the same time, pain and recovery change as the postoperative period progresses. The close relationship between time and exposure therefore partly arises from the clinical setting itself, although it also complicates their statistical separation.

Several features of the study support the present analyses. The standardized lumbar fusion cohort and randomized trial design provided a well-defined clinical setting. Detailed PCA event histories and pain assessments allowed subsequent PCA activity to be related to the pain state observed at the beginning of each interval without interpolation of pain scores. Plasma sampling at predefined postoperative time points allowed these observations to be related to oxycodone, S-ketamine, and metabolite exposure. These data permitted separate analyses of exposure build-up, longitudinal dosing activity, and the relationships between pain, PCA activity, and drug exposure.

Several limitations should be considered. Although the parent trial was randomized, the associations examined here are observational. Plasma concentrations were available at a limited number of postoperative sampling points. Finally, the analyses examined selected relationships within the PCA process rather than modelling pain, dosing behaviour, and drug exposure simultaneously.

In conclusion, our analyses suggest that during postoperative PCA, drug exposure develops with the patient’s dosing history while pain and recovery change over time. Current pain was associated with subsequent PCA activity, whereas the association between plasma oxycodone concentration and adequate analgesia weakened with postoperative time. Achieved S-ketamine exposure varied within randomized treatment groups and reflected the PCA dosing history as well as treatment allocation. These findings indicate that pain intensity, patient-controlled dosing behaviour, and drug exposure are interconnected but non-equivalent features of PCA.

## Data Availability

All data produced in the present study are available upon reasonable request to the authors

## Statements and Declarations

## Funding

This study was funded by State funding for university-level health research to Turku University Hospital (#13821 to TIS). The funder had no role in study design, conduct, data analysis, interpretation, manuscript preparation, or the decision to submit the manuscript for publication.

## Competing interests

The authors declare no competing interests.

## Ethics approval

The study protocol was reviewed and approved by the Ethics Committee of the Hospital District of Southwest Finland 103/1800/2016 and the Finnish National Agency for Medicines (KL no 135/2016).

## Consent to participate

All participants provided written informed consent before study participation.

## Patient consent for publication

Not applicable.

## AI disclosure

OpenAI’s ChatGPT was used to assist in manuscript editing, refinement of scientific presentation, and development and review of statistical analysis scripts. All generated content and code were reviewed, verified, and approved by the authors, who take full responsibility for the analyses and the manuscript.

## Acknowledgements

Maria Mattila, Aliisa Backman, and Daniel Backmansson were indispensable in handling the study samples. We thank Lisbet Partanen and Eija Mäkinen-Pulli for their skilful technical assistance. Eliisa Löyttyniemi, MSc, Institute of Biomedicine, University of Turku gave invaluable statistical help.

## Availability of data

Individual participant data are not publicly available because of restrictions related to patient privacy and the potential risk of re-identification. Further information may be obtained from the corresponding author, subject to applicable ethical and institutional approvals.

## Author contributions

TV contributed to study conduct, data analysis, statistical interpretation, and manuscript drafting and revision. ECVB contributed to study design, study conduct, data analysis, and manuscript drafting and revision. ME, MJK, JB, and AT contributed to bioanalytical analyses and manuscript revision. PT and VK contributed to study conduct, and manuscript revision. MP contributed to study design, data analysis, and manuscript drafting and revision. TIS contributed to study design, study conduct, data analysis, statistical interpretation, and manuscript drafting and revision. All authors critically reviewed and approved the final manuscript. TIS is the guarantor of the study.

## STROBE Statement—checklist of items that should be included in reports of observational studies

|  | Item No | Recommendation | Reported on page |
|---|---|---|---|
| **Title and abstract** | 1 | (*a*) Indicate the study’s design with a commonly used term in the title or the abstract | 1–2 |
|  |  | (*b*) Provide in the abstract an informative and balanced summary of what was done and what was found | 2 |
| **Introduction** |  |  |  |
| Background/rationale | 2 | Explain the scientific background and rationale for the investigation being reported | 4–5 |
| Objectives | 3 | State specific objectives, including any prespecified hypotheses | 5 |
| **Methods** |  |  |  |
| Study design | 4 | Present key elements of study design early in the paper | 6 |
| Setting | 5 | Describe the setting, locations, and relevant dates, including periods of recruitment, exposure, follow-up, and data collection | 6–8 |
| Participants | 6 | (*a*) *Cohort study*—Give the eligibility criteria, and the sources and methods of selection of participants. Describe methods of follow-up<br>*Case-control study*—Give the eligibility criteria, and the sources and methods of case ascertainment and control selection. Give the rationale for the choice of cases and controls<br>*Cross-sectional study*—Give the eligibility criteria, and the sources and methods of selection of participants | 6–7; ref. 12 |
|  |  | (*b*) *Cohort study*—For matched studies, give matching criteria and number of exposed and unexposed<br>*Case-control study*—For matched studies, give matching criteria and the number of controls per case | N/A |
| Variables | 7 | Clearly define all outcomes, exposures, predictors, potential confounders, and effect modifiers. Give diagnostic criteria, if applicable | 6–9 |
| Data sources/ measurement | 8* | For each variable of interest, give sources of data and details of methods of assessment (measurement). Describe comparability of assessment methods if there is more than one group | 6–9; Suppl. Methods |
| Bias | 9 | Describe any efforts to address potential sources of bias | 8–9, 17–18 |
| Study size | 10 | Explain how the study size was arrived at | 7 |
| Quantitative variables | 11 | Explain how quantitative variables were handled in the analyses. If applicable, describe which groupings were chosen and why | 8–9; Fig. 2 legend |
| Statistical methods | 12 | (*a*) Describe all statistical methods, including those used to control for confounding | 7–9 |
|  |  | (*b*) Describe any methods used to examine subgroups and interactions | 8–9 |
|  |  | (*c*) Explain how missing data were addressed | 7, 10; Suppl. Tables S3–S4* |
|  |  | (*d*) *Cohort study*—If applicable, explain how loss to follow-up was addressed<br>*Case-control study*—If applicable, explain how matching of cases and controls was addressed<br>*Cross-sectional study*—If applicable, describe analytical methods taking account of sampling strategy | 7, 10; Suppl. Fig. S1* |
|  |  | (*e*) Describe any sensitivity analyses | 8–9 |
| **Results** |  |  |  |
| Participants | 13* | (*a*) Report numbers of individuals at each stage of study—eg numbers potentially eligible, examined for eligibility, confirmed eligible, included in the study, completing follow-up, and analysed | 10–13; Suppl. Fig. S1 (p. 10); ref. 12 |
|  |  | (*b*) Give reasons for non-participation at each stage | 7, 10; Suppl. Fig. S1 (p. 10); ref. 12 |
|  |  | (*c*) Consider use of a flow diagram | Suppl. Fig. S1 (p. 10) |
| Descriptive data | 14* | (*a*) Give characteristics of study participants (eg demographic, clinical, social) and information on exposures and potential confounders | 10–12; Table 1; Figs. 1–2 |
|  |  | (*b*) Indicate number of participants with missing data for each variable of interest | 10; Table 2; Suppl. Tables S2–S5* |
|  |  | (*c*) *Cohort study*—Summarise follow-up time (eg, average and total amount) | 7–8 (24 h) |
| Outcome data | 15* | *Cohort study*—Report numbers of outcome events or summary measures over time<br>*Case-control study*—Report numbers in each exposure category, or summary measures of exposure<br>*Cross-sectional study*—Report numbers of outcome events or summary measures | 10–15; Figs. 1–2 |
| Main results | 16 | (*a*) Give unadjusted estimates and, if applicable, confounder-adjusted estimates and their precision (eg, 95% confidence interval). Make clear which confounders were adjusted for and why they were included | 8–9, 12–15; Table 2; Suppl. Tables S2–S5* |
|  |  | (*b*) Report category boundaries when continuous variables were categorized | Fig. 2 legend (visualization only) |
|  |  | (*c*) If relevant, consider translating estimates of relative risk into absolute risk for a meaningful time period | Fig. 3C (predicted probabilities) |
| Other analyses | 17 | Report other analyses done—eg analyses of subgroups and interactions, and sensitivity analyses | 8–9, 12–15; Suppl. Tables S3–S5* |
| **Discussion** |  |  |  |
| Key results | 18 | Summarise key results with reference to study objectives | 16 |
| Limitations | 19 | Discuss limitations of the study, taking into account sources of potential bias or imprecision. Discuss both direction and magnitude of any potential bias | 17–18 |
| Interpretation | 20 | Give a cautious overall interpretation of results considering objectives, limitations, multiplicity of analyses, results from similar studies, and other relevant evidence | 16–19 |
| Generalisability | 21 | Discuss the generalisability (external validity) of the study results | 18 (clinical setting; limited discussion)* |
| **Other information** |  |  |  |
| Funding | 22 | Give the source of funding and the role of the funders for the present study and, if applicable, for the original study on which the present article is based | 20 |

\*Give information separately for cases and controls in case-control studies and, if applicable, for exposed and unexposed groups in cohort and cross-sectional studies.
Note: An Explanation and Elaboration article discusses each checklist item and gives methodological background and published examples of transparent reporting. The STROBE checklist is best used in conjunction with this article (freely available on the Web sites of PLoS Medicine at http://www.plosmedicine.org/, Annals of Internal Medicine at http://www.annals.org/, and Epidemiology at http://www.epidem.com/). Information on the STROBE Initiative is available at www.strobe-statement.org.
Audit notes (\*): Items 12c and 14b: analysis-specific sample sizes are reported, not missing counts for each variable; the BLQ substitution rule is not stated in the inspected manuscript. Items 12d and 13a–b: final participant-flow counts and exclusion reasons must be checked against the final Supplementary Figure S1 and parent trial report (reference 12). Item 16a: adjusted estimates are reported; unadjusted estimates are not tabulated. Item 17: quantified-only analyses are described in Methods, but their numerical results are not reported in the inspected manuscript or supplement. Item 21: the clinical setting is described; external validity is not explicitly discussed.
Page references are based on DRK\_ms3\_201126\_v2.docx (latest manuscript available for inspection); check pagination against the final Word file before submission.

## SUPPLEMENTARY MATERIAL

## 1. Quantitative HPLC-MS/MS analysis of oxycodone, noroxycodone, oxymorphone and noroxymorphone in human plasma

### Chemicals and supplies

Oxycodone, noroxycodone, oxymorphone, noroxymorphone, oxycodone-D_3_, noroxycodone-D_3_ and oxymorphone-D_3_ were purchased from Sigma-Aldrich (St. Louis, MO, USA). LC-MS-grade methanol was purchased from Honeywell / Riedel-de Haën (Seelze, Germany), and LC-MS grade formic acid was purchased from VWR International (Fontenay-Sous-Bois, Paris, France). Ammonium acetate and 25% ammonia solution were purchased from Merck (Darmstadt, Germany). Water used was purified with Millipore Synergy UV (Merck KGaA, Darmstadt, Germany) system.

### Preparation of calibration standards and quality control samples

Stock solutions and working solutions were prepared in methanol. Working solutions contained either all four analytes or all three internal standards. Calibration standards and quality control (QC) samples were prepared by spiking pooled, drug-free human K_2_EDTA plasma with appropriate volumes of working solutions. The calibration standards contained oxycodone, noroxycodone, oxymorphone, and noroxymorphone at 0.05, 0.1, 0.3, 2.0, 10.0, 20.0, 45.0, and 50.0 ng/mL. QC concentrations were 0.15, 12.0, and 40.0 ng/mL. For oxycodone, additional dilution controls of 200 ng/mL (10× dilution) and 2000 ng/mL (100× dilution) were prepared for selected batches. All calibration standards, QC samples, and study samples were stored at –70 °C until analysis and processed identically.

### Sample preparation

Study samples, calibration standards, and QC samples were prepared using solid-phase extraction (SPE). Briefly, 200 µL of plasma was mixed with 25 µL of internal standard working solution (100 ng/mL) and 775 µL of 0.1 % formic acid in water (v/v). After vortex-mixing, samples were transferred to conditioned Sep-Pak® tC18 96-well plates (Waters, Milford, MA, USA). Analytes were eluted with 1 mL methanol, evaporated under nitrogen, and reconstituted in 100 µL of 20 % methanol in water (v/v). The extracts were transferred to autosampler vials, centrifuged at 7500×g for 2 min, and subjected to High-performance liquid chromatography–tandem mass spectrometry (HPLC–MS/MS) analysis.

### HPLC-MS/MS analysis and quantitation

The HPLC-MS/MS system consisted of a Sciex ExionLC™ AD HPLC and a Sciex QTrap 6500^+^ triple quadrupole mass spectrometer (AB Sciex LP, Concord, ON, Canada). Chromatographic separation was performed with a Kinetex™ EVO C18 analytical column (50 × 4.6 mm, particle size 5 µm) coupled with an EVO C18 guard column from Phenomenex (Torrance, CA, USA). The mobile phase consisted of (A) 5 mM ammonium acetate in water, adjusted to pH 9.4, and (B) 100 % methanol, delivered at a constant flow rate of 0.45 mL/min. The gradient was as follows: 0.0–1.0 min, 10 % B; 1.0–2.0 min, linear increase to 40 % B; 2.0–5.0 min, 40 % B; 5.0–6.0 min, linear increase to 95 % B; 6.0–8.0 min, 95 % B; 8.0–8.1 min, linear decrease to 10 % B; and 8.1–10.0 min, 10 % B. The injection volume was 10 µL. The column oven temperature was 50 °C. Mass spectrometric detection was carried out using positive Turbo Ion Spray (TIS) ionization and selected reaction monitoring (SRM) mode. The SRM transitions and ion-specific parameters are summarized in Supplementary Table S1. The ion source temperature was 500 °C. The nebulizer gas (Gas 1) and turbo gas (Gas 2) settings were 50 psi. Curtain gas (nitrogen) was set to 20 psi. The TIS voltage setting was 4500 V. Dwell time was 250 ms for all ion transitions. Quantitation was based on analyte-to-internal standard peak area ratios. Chromatograms were acquired and processed using Analyst® software (version 1.6.3, AB Sciex). Calibration curves were constructed using weighted (1/x²) linear regression.

**Supplementary Table S1.**
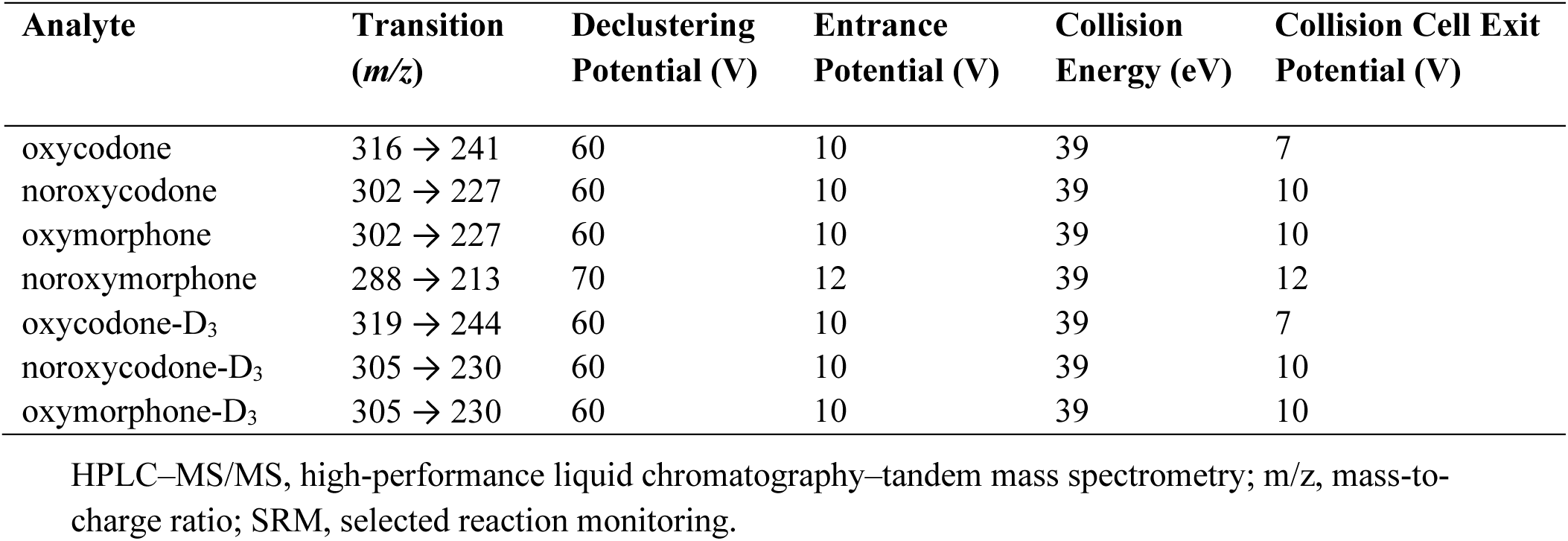
SRM transitions and ion-specific parameters used for HPLC–MS/MS quantitation.

### Method validation and analytical performance

Fit-for-purpose method validation was conducted for oxycodone, noroxycodone, oxymorphone and noroxymorphone in human K_2_EDTA plasma prior to sample analysis. Method performance was considered acceptable if the back-calculated concentrations of the calibration standards were within ±15 % of the nominal value (± 20 % at lower limit of quantification, LLOQ). In addition, the mean concentration of the quality control samples had to be within ±15 % of the nominal concentration at low, mid and high validation QC levels and within ± 20 % at LLOQ. The precision (CV (%)) had to be ≤ 15 % at low, mid and high validation QC levels and ≤ 20 % at LLOQ.

In fit-for-purpose validation, the linear range for all four analytes was 0.05 ng/mL – 50 ng/mL (8-point calibration curves). Intra-assay accuracies of six parallel quality control (QC) samples at four different concentration levels (and additional dilution QC samples for oxycodone) were 93.1 % – 105 %, 97.1 % – 105 %, 94.5 % – 104 %, and 90.9 % – 100 % for oxycodone, noroxycodone, oxymorphone, and noroxymorphone, respectively. The corresponding intra-assay coefficients of variation were 1.03 % – 5.68 % (oxycodone), 1.73 % – 7.58 % (noroxycodone), 1.40 % – 13.2 % (oxymorphone), and 3.94 % – 12.2 % (noroxymorphone).

For the study sample analysis, the back-calculated concentrations of the calibration standards had to be within ±15 % of the nominal values (± 20 % at LLOQ) and at least 6 of 8 calibration standards were required to meet this criterion. The accuracy of the QC samples had to be ± 15% of the nominal values and at least 50 % at each concentration level and at least four of the six QC samples had to fulfill the criterion. The concentration results were acquired in nine analysis batches for noroxycodone, oxymorphone and noroxymorphone. Concentration result for oxycodone was acquired in eleven batches. The inter-assay accuracy of the QC samples was 92.6 % – 108 % for oxycodone, 95.6 % – 100 % for noroxycodone, 98.0 % – 105 % for oxymorphone and 104 % for noroxymorphone. The corresponding inter-assay coefficients of variation were 1.55 % – 4.86 %, 3.37 % – 3.91 %, 3.37 % – 5.28 %, and 3.81 % – 6.87 %, respectively.

## 2. Determination of ketamine, norketamine and 6-hydroxynorketamine concentrations in human plasma

### Chemicals and supplies

The analytes ketamine and norketamine were purchased from Merck (Darmstadt, Germany), and 6-hydroxynorketamine from Bio-Techne (Abingdon, UK), and the deuterated internal standards (S)-ketamine-d6 and norketamine-d4 from LGC (Guildford, UK).

### Sample preparation

The spiked plasma calibrators and quality control samples, and the patient plasma samples, were pretreated using Oasis MCX µelution 96-well plates (Waters, Milford, MA, USA). The solid phase extraction (SPE) was carried out with vacuum manifold and according to the manufacturer’s instructions. Briefly, the SPE plate was conditioned with 200 µL of methanol followed by 200 µL of water. In a separate deep well polypropylene plate, aliquots of the plasma samples were mixed with an equal volume of internal standards containing 4% orthophosphoric acid solution. Next, 270 µL of the mixture was loaded into the SPE plate, which was first washed with 200 µL of 2% formic acid, and then with 200 µL of methanol. After the washes, the plate was dried under vacuum for 1 minute. The analytes were eluted from the SPE plate into polypropylene 96-well plate with two 50 µL aliquots of 3% ammonium hydroxide in 60% acetonitrile: 40% methanol. The sample extracts were evaporated to dryness under vacuum at 58 ⁰C, using a Genevac® miVac centrifugal concentrator (Thermo Fisher Scientific, Waltham, MA, USA), and reconstituted in 50 µL of water.

### HPLC-MS/MS analysis and quantitation

The determination of the analytes was carried out using Sciex Exion liquid chromatography and Sciex 6500 QTRAP mass spectrometer interfaced with an electrospray ion source (AB Sciex, Toronto, ON, Canada). Analytes were separated using an Atlantis T3 column, 100 × 2.1 mm, 3 µm particle size (Waters, Milford, MA, USA) with 0.1% formic acid as a mobile phase A and acetonitrile as mobile phase B at flow rate 0.3 mL/min. The gradient programme for the mobile phase B was a linear gradient from 5% to 30% over 4.5 min, followed by rapid increase to 90% in 0.1 min, after which the mobile phase B was kept constant at 90% for 2 min, followed by balancing at 5% before the next injection. The mass spectrometer was operated in positive electrospray ionization mode (ESI+) and the mass-to-charge (m/z) ion transitions were 238 → 125 for ketamine, 224 → 125 for norketamine and 240 → 125 for 6-hydroxynorketamine. Corresponding deuterium-labelled compounds were used as internal standards except for 6-hydroxynorketamine, which was quantified with norketamine-d4 as internal standard. The lower limits of quantification were 0.025 ng/mL for ketamine, 0.25 ng/mL for norketamine, and 0.5 ng/mL for 6-hydroxynorketamine. Between-day precision (CV) was below 15%, and accuracy was within ±15%, except at the LLOQ, where precision and accuracy met the corresponding 20% acceptance limits.

**Supplementary Table S2.**
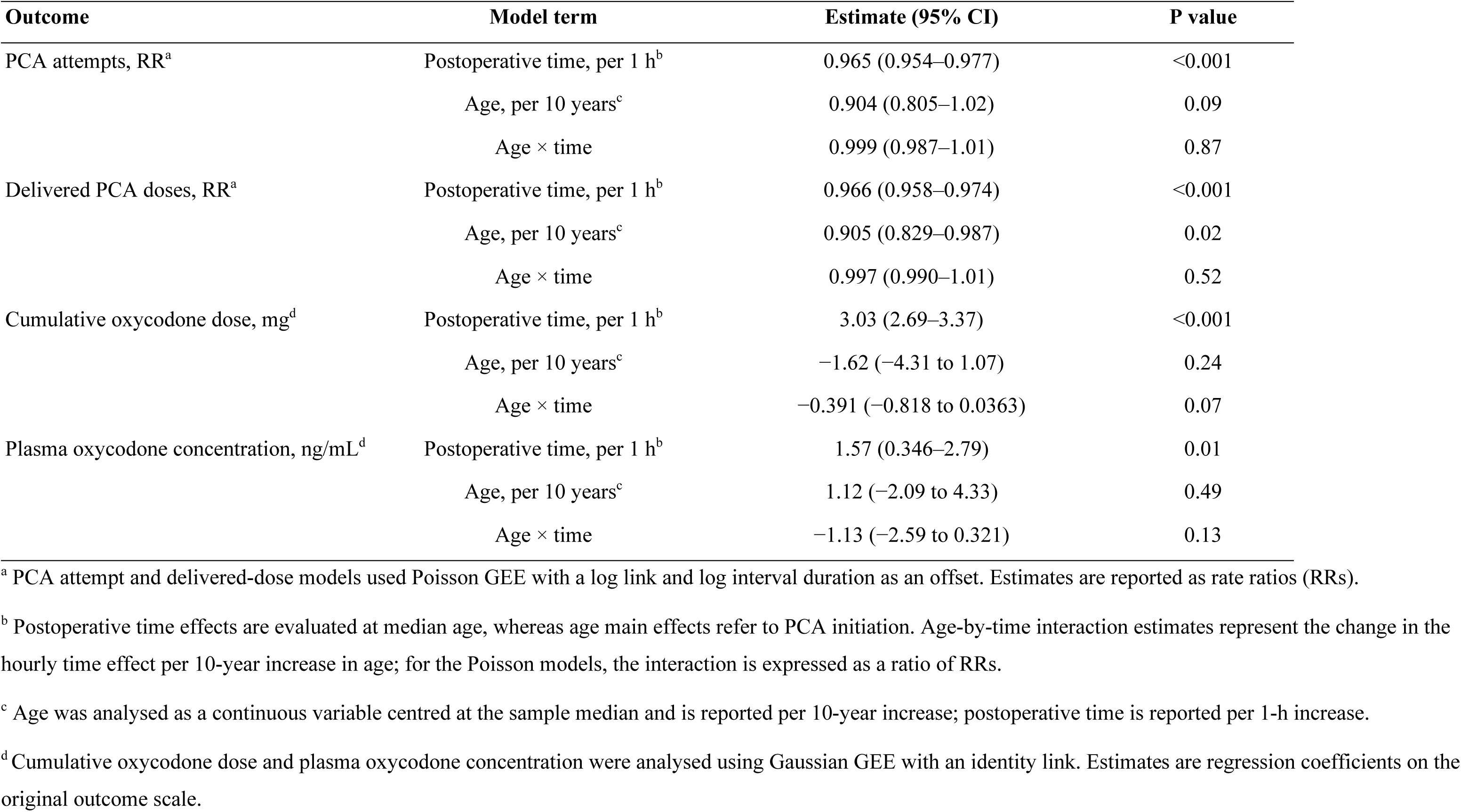
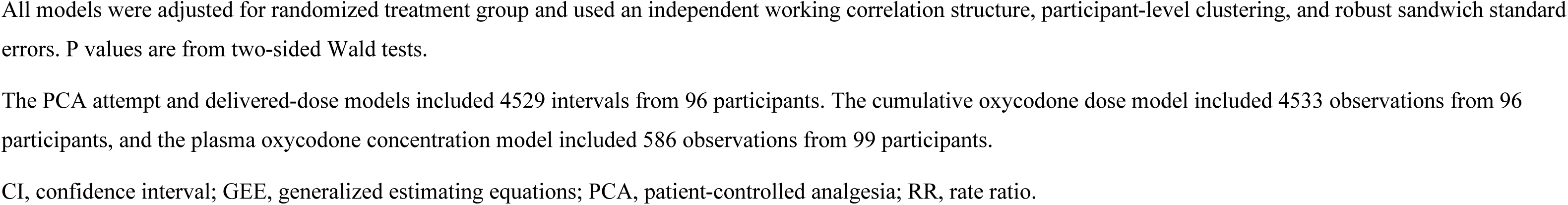
Longitudinal associations of postoperative time and age with PCA activity, cumulative oxycodone dose, and plasma oxycodone concentration during the first 24 h after PCA initiation.

**Supplementary Table S3.** Sensitivity of pain–PCA and oxycodone–analgesia associations to adjustment for measured S-ketamine and metabolite concentrations.

| **Outcome<sup>a</sup>** | **Without S-ketamine or metabolites (M0)** | **+ S-ketamine (M1)** | **+ S-ketamine and metabolites (M2)** |
|---|---|---|---|
| Subsequent PCA attempts | 1.08 (0.976–1.19) | 1.08 (0.982–1.20) | 1.09 (0.987–1.20) |
| Subsequent delivered doses | 1.06 (1.00–1.12) | 1.06 (1.01–1.13) | 1.07 (1.01–1.13) |
| Adequate analgesia: oxycodone × time, P | 0.004 | 0.002 | 0.003 |

<sup>a</sup> For the two PCA outcomes, estimates are rate ratios (RRs) with 95% confidence intervals (CIs) per 1-point increase in current pain intensity at rest (NRSR). For adequate analgesia, values are P values from two-sided Wald tests of the plasma oxycodone concentration-by-time interaction.
Analyses were restricted to participants randomized to S-ketamine-containing PCA (G2–G4). Models M0–M2 were fitted to identical complete-case subsets within each outcome, retaining the predefined substitution values for concentrations below the lower limit of quantification. The PCA activity models included 194 intervals from 71 participants, and the adequate analgesia models included 261 matched concentration–NRSR observations from 71 participants.
All models used an exchangeable working correlation structure, participant-level clustering, and robust sandwich standard errors.
M0, reference model without adjustment for measured S-ketamine or metabolite concentrations; M1, M0 additionally adjusted for S-ketamine concentration; M2, M0 additionally adjusted for S-ketamine, norketamine, and 6-hydroxynorketamine concentrations. All three analyte concentrations were log<sub>2</sub>-transformed and centred within randomized treatment group and sampling epoch.
CI, confidence interval; GEE, generalized estimating equations; NRSR, numerical rating scale pain score at rest; PCA, patient-controlled analgesia; RR, rate ratio.

**Supplementary Table S4.** Associations of measured S-ketamine and metabolite concentrations with adequate analgesia across all analysed and later sampling epochs.

| **Analysis** | **S-ketamine<sup>a</sup>** | **Norketamine<sup>a</sup>** | **6-hydroxynorketamine<sup>a</sup>** | **Joint P-value<sup>b</sup>** |
|---|---|---|---|---|
| All analysed epochs, 15–1440 min | 0.691 (0.517–0.924) | 1.15 (0.848–1.55) | 0.963 (0.822–1.13) | 0.09 |
| Later epochs, 60–1440 min | 0.862 (0.582–1.28) | 1.06 (0.642–1.74) | 0.937 (0.675–1.30) | 0.79 |

<sup>a</sup> Estimates are odds ratios (ORs) with 95% confidence intervals (CIs) for adequate analgesia per doubling of plasma analyte concentration. Concentrations were log<sub>2</sub>-transformed and centred within randomized treatment group and sampling epoch.
<sup>b</sup> P values are from 3-df Wald tests of the joint contribution of S-ketamine, norketamine, and 6-hydroxynorketamine concentrations.
All estimates were derived from logistic GEE models (M2) including the three analyte concentrations simultaneously, together with plasma oxycodone concentration, postoperative time, their interaction, age, and randomized treatment group. Models used an exchangeable working correlation structure, participant-level clustering, and robust sandwich standard errors.
Analyses were restricted to participants randomized to S-ketamine-containing PCA (G2–G4), retaining the predefined substitution values for concentrations below the lower limit of quantification. The analysis including all four sampling epochs (15, 60, 240, and 1440 min) comprised 261 matched concentration–NRSR observations from 71 participants. The later-epoch analysis (60, 240, and 1440 min) comprised 198 observations from 71 participants.
CI, confidence interval; df, degrees of freedom; GEE, generalized estimating equations; NRSR, numerical rating scale pain score at rest; OR, odds ratio; PCA, patient-controlled analgesia.

**Supplementary Table S5.** Exploratory longitudinal associations of body weight with PCA activity, cumulative oxycodone dose, and plasma oxycodone concentration during the first 24 h after PCA initiation.

| **Outcome** | **Weight, per 10 kg, estimate (95% CI)** | **P** | **Weight × time estimate (95% CI)** | **P** |
|---|---|---|---|---|
| PCA attempts, RR<sup>a</sup> | 1.02 (0.949–1.09) | 0.60 | 1.00 (0.996–1.01) | 0.31 |
| Delivered doses, RR<sup>a</sup> | 1.04 (0.987–1.10) | 0.14 | 1.00 (0.996–1.01) | 0.60 |
| Cumulative oxycodone, mg<sup>b</sup> | 0.633 (−0.784 to 2.05) | 0.38 | 0.0922 (−0.143 to 0.327) | 0.44 |
| Plasma oxycodone concentration, mg/mL<sup>b</sup> | −1.44 (−3.14 to 0.254) | 0.10 | 0.365 (−0.397 to 1.13) | 0.35 |

Body weight replaced age in these exploratory models.
<sup>a</sup> PCA attempt and delivered-dose models used Poisson GEE with a log link and log interval duration as an offset. Estimates are reported as rate ratios (RRs).
<sup>b</sup> Cumulative oxycodone dose and plasma oxycodone concentration were analysed using Gaussian GEE with an identity link. Estimates are regression coefficients on the original outcome scale.
Body weight was analysed as a continuous variable centred at the sample median and is reported per 10-kg increase. Weight main effects refer to PCA initiation. Weight-by-time interaction estimates represent the change in the hourly time effect per 10-kg increase in body weight; for the Poisson models, the interaction is expressed as a ratio of RRs.
All models were adjusted for randomized treatment group and used an independent working correlation structure, participant-level clustering, and robust sandwich standard errors. P values are from two-sided Wald tests.
The PCA attempt and delivered-dose models included 4529 intervals from 96 participants. The cumulative oxycodone dose model included 4533 observations from 96 participants, and the plasma oxycodone concentration model included 586 observations from 99 participants.
CI, confidence interval; GEE, generalized estimating equations; PCA, patient-controlled analgesia; RR, rate ratio.

**Supplementary Figure S1:**
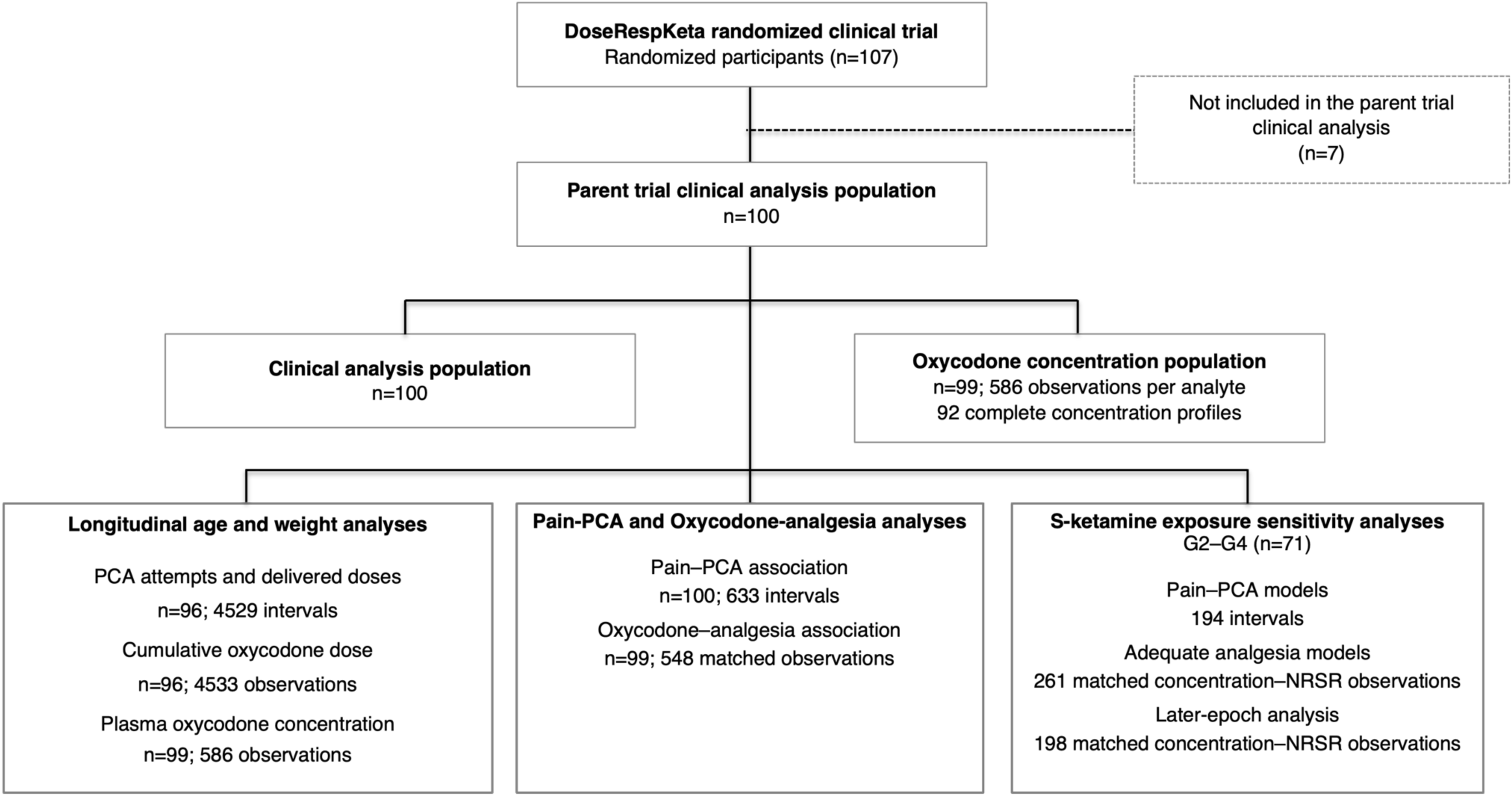
Participant flow and analysis populations for the present secondary analysis. Flow diagram showing derivation of the analysis populations from the DoseRespKeta trial and the sample sizes used in the principal analyses. Analysis populations overlap and are not mutually exclusive. Observation counts reflect the data available for each analysis. NRSR, numerical rating scale pain score at rest; PCA, patient-controlled analgesia.

**Supplementary Figure S2.**
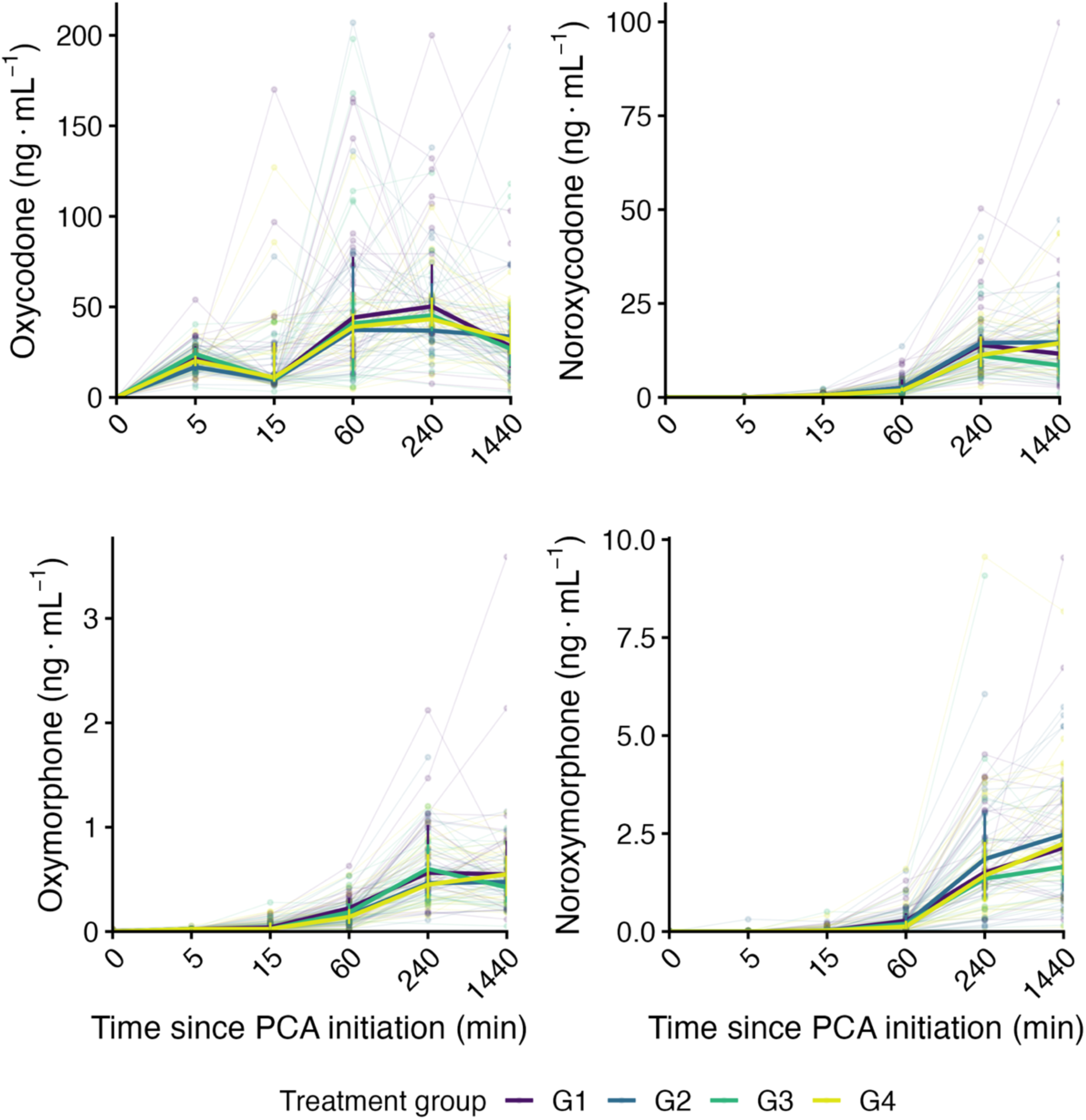
Plasma concentration profiles of oxycodone, noroxycodone, oxymorphone, and noroxymorphone by randomized treatment group. Grey lines and points represent individual concentration trajectories and observations; coloured lines and ribbons indicate medians and interquartile ranges. Protocol sampling epochs are displayed at equal spacing. All groups received oxycodone 1 mg/mL, with S-ketamine concentrations of 0 (G1), 0.25 (G2), 0.5 (G3), and 0.75 mg/mL (G4) in the PCA solution. The three highest oxycodone concentrations at 24 h were omitted from the display of individual observations to improve visualization but were retained in all summary calculations and analyses. PCA, patient-controlled analgesia.

**Supplementary Figure S3.**
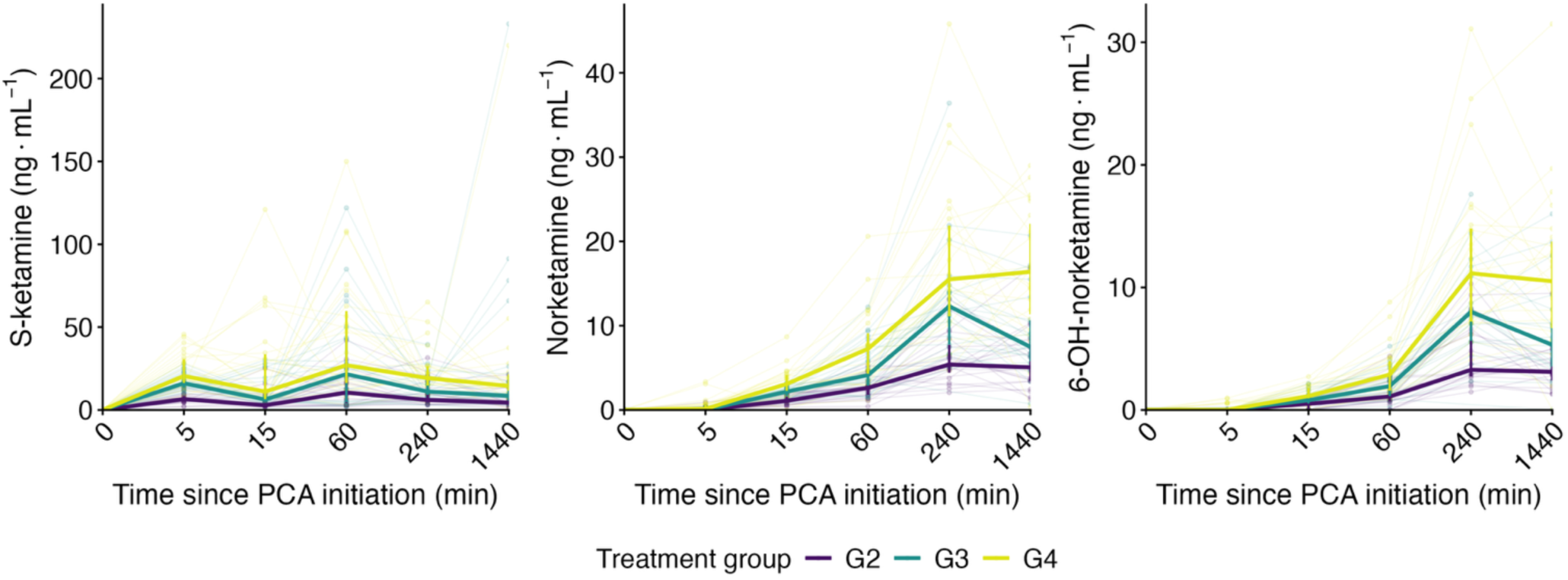
Plasma concentration profiles of S-ketamine, norketamine, and 6-hydroxynorketamine by randomized treatment group. Grey lines and points represent individual concentration trajectories and observations; coloured lines and ribbons indicate medians and interquartile ranges. Protocol sampling epochs are displayed at equal spacing. Groups G2, G3, and G4 received PCA solutions containing oxycodone 1 mg/mL and S-ketamine 0.25, 0.5, and 0.75 mg/mL, respectively. PCA, patient-controlled analgesia.

## Notes

### Competing Interest Statement

The authors have declared no competing interest.

### Clinical Trial

NCT02994173

### Author Declarations

Institutional review board of of the Hospital District of Southwest Finland gave ethical approval for this work (103/1800/2016)

### Summary of Updates

Manuscript and Supplemental files updated.

